# MISFOLDING OF ALPHA-SYNUCLEIN AS BLOOD-BASED BIOMARKER FOR PARKINSON’S DISEASE

**DOI:** 10.64898/2025.12.19.25342662

**Authors:** Lennart Langenhoff, Jonas Simon, Sandrina Weber, Diana Hubert, Martin Schuler, Marvin Mann, Vuk Puzovic, Grischa Gerwert, Adrian Höveler, Léon Beyer, Lars Tönges, Carsten Kötting, Jörn Güldenhaupt, Brit Mollenhauer, Klaus Gerwert

## Abstract

Parkinson’s disease (PD), dementia with Lewy bodies (DLB) and multiple system atrophy (MSA) are characterized by pathological aggregation of misfolded alpha-synuclein (αSyn) in the central and peripheral nervous systems. Seed amplification assays (SAAs) can detect misfolded αSyn in cerebrospinal fluid (CSF), allowing a more precise biological classification of these diseases. Translating biomarker-based diagnostics to blood represents a crucial milestone to democratize access to molecular testing, as blood sampling is minimally invasive and widely scalable. However, current blood-based SAA approaches require extensive sample preprocessing, which introduces variability and has yielded inconsistent results. Here, we present the immuno-infrared-sensor (iRS) platform, which detects misfolded αSyn directly in blood serum without sample preprocessing and without amplification. The iRS measures the secondary-structure distribution of αSyn by distinguishing α-helical/random-coil monomers from β-sheet-rich oligomers and fibrils. This conformational distribution reflects disease-associated structural changes in blood and provides information beyond the binary readout of SAAs. In a combined cohort (n = 127) consisting of 74 individuals with synucleinopathies (PD, DLB, MSA and isolated rapid eye movement sleep behavior disorder (iRBD)) and 53 controls, the assay achieved a receiver operating characteristic area under the curve (ROC AUC) of 0.94 (95% confidence interval (CI), 0.88–0.99), with 88% sensitivity and 89% specificity in serum using a dual-threshold classification that categorized individuals as controls (no misfolding, green), at risk (low misfolding, yellow) or diseased (high misfolding, red). By extending the iRS platform from CSF to blood serum, this study demonstrates αSyn misfolding as a promising blood-based structural biomarker for synucleinopathies. The iRS assay provides a broadly applicable platform technology for more precise biological classification, disease monitoring and assessment of drug response for PD in peripheral blood.

---

Subject Categories: Biomarkers Neuroscience

## Introduction

After Alzheimer’s disease (AD), Parkinson’s disease (PD) is the second most common progressive neurodegenerative disorder, characterized by a spectrum of motor and non-motor symptoms that together impose a substantial burden on individuals and public health^1,2^. Current clinical diagnostic criteria depend on the manifestation of characteristic motor features, including bradykinesia or akinesia, rigidity, resting tremor and postural instability, at a stage when a substantial loss of dopaminergic neurons in the substantia nigra has already occurred^1,3,4^. Alpha-synuclein (αSyn) pathology, particularly the misfolding and aggregation of αSyn into oligomers, fibrils and Lewy body deposits, is a neuropathological hallmark not only of PD, the most common synucleinopathy, but also of multiple system atrophy (MSA) and dementia with Lewy bodies (DLB)^5^. Consequently, assays that detect aggregated or misfolded αSyn in biofluids have emerged as leading biomarker approaches as the field advances toward a more biologically anchored disease definition^6–9^.

In cerebrospinal fluid (CSF), αSyn seed amplification assays (SAAs) have demonstrated high diagnostic accuracy for detecting pathological αSyn conformers in clinically diagnosed PD and prodromal synucleinopathy patients^10^, supporting the concept that proteopathic seeds in body fluids reflect central nervous system pathology^9,11,12^. Nevertheless, CSF collection is invasive, requires specific training and is not well-suited to large-scale screening or frequent longitudinal monitoring^13^, while SAAs face challenges in standardization and depend strongly on assay conditions and substrate quality^14,15^. In the search for less invasive biomarkers, skin biopsies have shown diagnostic utility by detecting misfolded αSyn aggregates in patients with PD and other synucleinopathies, with studies reporting high sensitivity and near-perfect specificity^16,17^. However, the procedure remains invasive and shows reduced sensitivity at early disease stages, limiting broader clinical implementation and highlighting the need for biomarkers derived from more accessible sample matrices^18–20^. This need is particularly urgent for screening individuals at elevated risk of developing PD in anticipation of upcoming prevention trials.

Blood serum is widely recognized as an attractive alternative sample matrix for PD biomarkers because it is minimally invasive, easily accessible and suitable for repeated sampling and established routine processing in longitudinal studies and population screening^21^. Translating αSyn detection from CSF to blood-based matrices is challenging, as serum and plasma contain substantial peripheral αSyn contributions, variable αSyn concentrations and a complex protein matrix that complicates precise detection of distinct αSyn conformers^21,22^. Recently, two blood-based SAAs for detecting misfolded αSyn have shown promising results in distinguishing individuals with PD from controls. One approach enriches αSyn directly from serum using immunoprecipitation^23^, while the other isolates blood-derived neuronal extracellular vesicles. However, both rely on sensitive and error-prone preprocessing steps, raising concerns about assay robustness and suitability for routine clinical application^14,28^.

The immuno-infrared-sensor (iRS) is a label-free spectroscopic approach that measures the secondary structure distribution of immunocaptured proteins (*Extended Data Fig. 1*). In the iRS platform, αSyn is captured on the biofunctionalized surface of the internal reflection element (IRE) used for attenuated total reflection (ATR) by specific antibodies. The infrared amide I band of αSyn is then recorded using Fourier transform infrared (FTIR) spectroscopy. The amide I band position typically ranges from about 1650 cm^-1^, dominated by α-helical or random-coil monomers, to around 1630 cm^-1^ when β-sheet enriched species are more abundant. A shift in amide I absorbance toward lower wavenumbers therefore reflects increasing αSyn misfolding within the sample. Difference spectroscopy is used to subtract background signals and isolate the αSyn-specific absorbance. By quantifying the relative abundance of β-sheet versus α-helical or random-coil conformers, the iRS technique provides a direct readout of protein misfolding without amplification steps or sample preprocessing. The iRS platform has already demonstrated excellent diagnostic performance in detecting misfolded amyloid-beta (Aβ) in CSF and blood plasma across different AD cohorts, including in individuals up to 17 years before the onset of clinical symptoms^29–34^. More recently, we extended the iRS assay to synucleinopathies, showing that αSyn misfolding in CSF distinguishes individuals with synucleinopathies from controls with high diagnostic accuracy^35^.

Here, we expanded the iRS platform to blood serum from routine collection and evaluate its potential as a biomarker for PD, using a serum-optimized biochip surface specifically designed to overcome challenges associated with complex protein matrices. We analyzed serum samples from clinically diagnosed synucleinopathies, including PD, MSA, DLB and isolated rapid eye movement sleep behavior disorder (iRBD), as well as controls, using the iRS platform across a discovery and an independent validation cohort. Diagnostic performance metrics such as receiver operating characteristic area under the curve (ROC AUC), sensitivity, specificity and P values were evaluated and compared with established amplification-based approaches. Our findings show that direct, label-free assessment of αSyn misfolding in serum allows noninvasive detection of synucleinopathies and may support a biologically anchored diagnosis in clinical practice.

## Results

### Sensor surface characterization and controls

The iRS platform has previously demonstrated that αSyn misfolding in CSF serves as a promising fluid biomarker for PD^35^. However, adapting the assay from CSF to blood serum required extensive optimization of the biochip to overcome serum-specific challenges. Briefly, the sensor surface charge was chemically adjusted to achieve a neutral surface charge, minimizing nonspecific protein adsorption. Additionally, the capture molecule was changed from a full-length immunoglobulin G (IgG) antibody, previously used for CSF measurements, to a single-chain variable fragment (scFv), thereby avoiding Fc-mediated off-target binding and improving specificity for αSyn in serum. The performance of the sensor surface and capture antibody was evaluated using iRS control measurements in combination with orthogonal indirect enzyme-linked immunosorbent assays (ELISA) to quantify αSyn reduction. Further details will become publicly available upon patent publication in 2026.

### Surface inertness and specificity

The newly developed biochip surface for blood serum measurements was first evaluated for nonspecific binding by assessing surface inertness in the absence of the capture antibody, using excessive amounts of αSyn conformers at concentrations far exceeding physiological serum levels (*Extended Data Fig. 2*). To further challenge the surface, human serum albumin (HSA) at a concentration of 50 mg/ml was tested for nonspecific adsorption (*Extended Data Fig. 2*). No detectable nonspecific binding of either αSyn conformers or HSA was observed, confirming high blocking efficiency and surface inertness of the chemical-functionalized biochip without antibodies. To assess surface performance under more physiologically relevant assay conditions, measurements were performed on capture-functionalized surfaces using the certified serum reference material (Serodos^®^), which contains analyte concentrations within the normal physiological range but lacks αSyn. When Serodos^®^ was spiked with 20 ng of either monomeric αSyn or αSyn preformed fibrils (PFFs), the amide I center of mass (CoM) shifted from 1641.5 cm^-1^ (monomer) to 1639.3 cm^-1^ (PFFs), indicating that the iRS readout discriminates between distinct αSyn conformations under physiologically relevant conditions (*Fig. 1*). The CoM provides a more robust readout than the absolute absorbance band maximum, as it is less affected by water vapor artifacts and noise and was therefore used for initial analyses. To evaluate αSyn binding efficiency on the iRS platform, αSyn concentrations were quantified using an indirect αSyn ELISA before and after applying a representative serum sample to sensor surfaces functionalized with either an active antibody fragment (wild-type, WT) or a binding-deficient dead mutant^36^ (DM) serving as a negative control (*Extended Data Fig. 3*). The exemplary original sample contained 9.9 ng/mL of αSyn. After measurement, the supernatant from the active antibody surface retained 3.3 ng/mL, indicating approximately 67% specific αSyn binding. In contrast, the DM surface showed a minimal reduction to 8.8 ng/mL, suggesting only 11% nonspecific loss, likely associated with system volume effects.

**Fig. 1:**
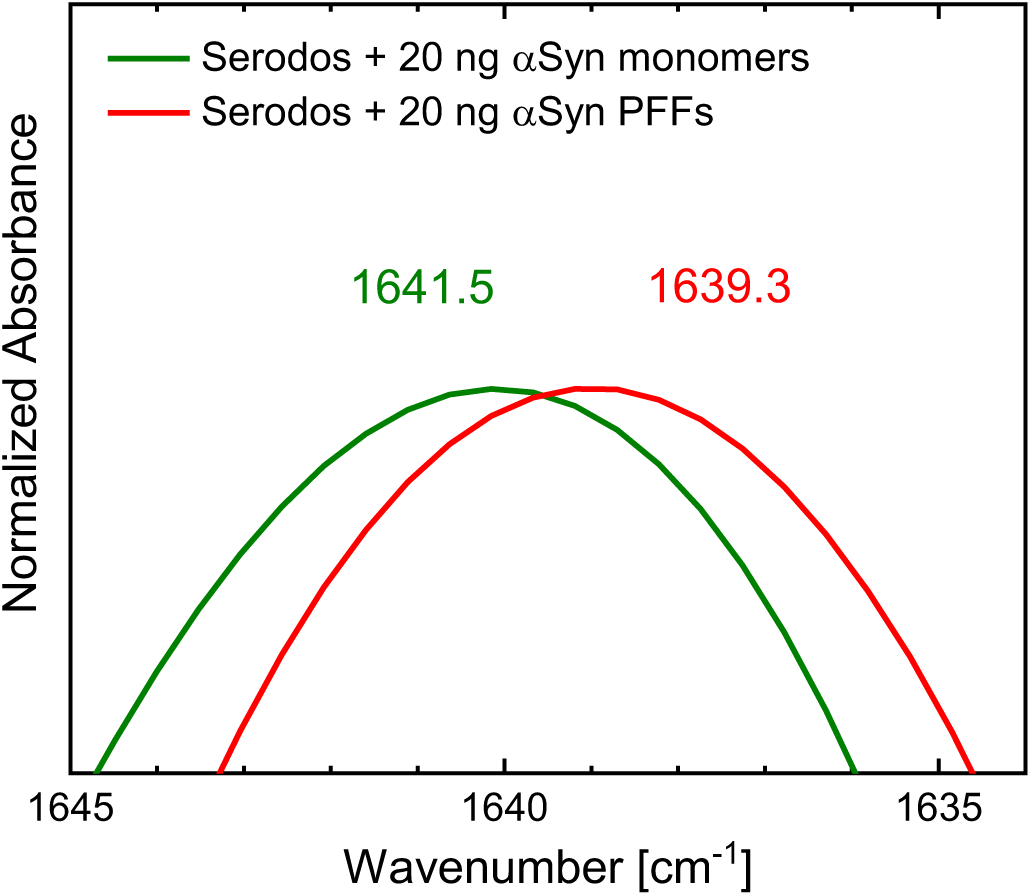
Specific iRS detection of monomeric and fibrillar αSyn in serum reference material. Zoom on amide I absorbance spectra of certified serum reference material (Serodos^®^, 300 µl) spiked with 20 ng of αSyn, either as monomer (green) or PFF (red), measured on active antibody-functionalized iRS surfaces. The amide I CoM is 1641.5 cm^-1^ for monomeric αSyn and 1639.3 cm^-1^ for PFFs, revealing distinct secondary-structure conformations in complex matrices. Abbreviations: αSyn alpha-synuclein, CoM center of mass, iRS immuno-infrared-sensor, PFFs preformed fibrils

### Transfer from CSF to blood serum

To translate the iRS platform technology from CSF to blood serum, the biochip surface chemistry was optimized to address the increased matrix complexity of blood. The modified surface was tailored for serum measurements and evaluated against the established surface for CSF measurements using paired CSF and serum samples from the same individuals, including normal pressure hydrocephalus (NPH) patients without PD (n = 5) and PD/DLB patients (n = 3) as shown in *Fig. 2*. Both CSF and serum analyses showed that misfolded αSyn cases (PD/DLB) exhibited lower amide I CoM wavenumbers compared with control samples. Serum amide I CoM values were consistently shifted by about 1.0 cm^-1^ toward higher wavenumbers, suggesting a matrix-induced assay shift or different αSyn distributions for serum measurements. Despite the limited sample size, the comparable serum/CSF trends encouraged further serum analysis in larger discovery and validation cohorts.

**Fig. 2:**
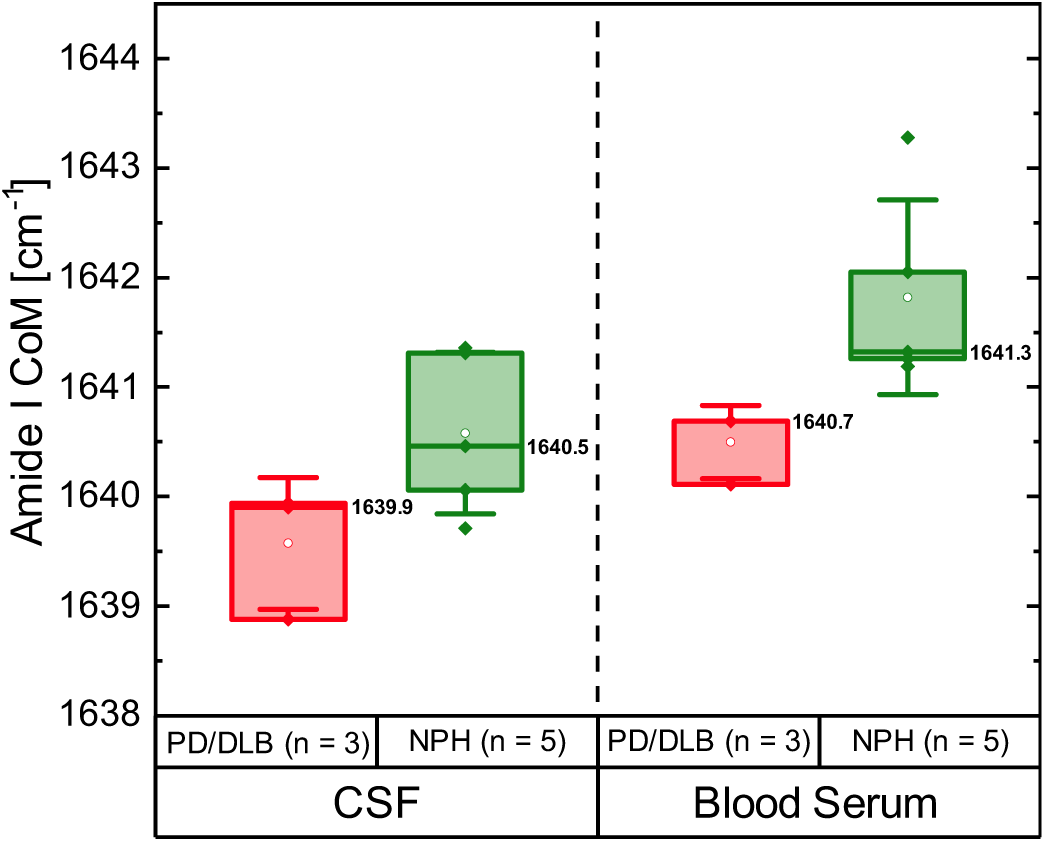
Paired CSF and serum comparison of αSyn secondary structure distribution for assay validation in NPH without PD and PD/DLB cases using the iRS platform. Boxplots of the amide I CoM (upper 10–20% of the amide I band) are shown for NPH cases without PD (controls, n = 5) and PD/DLB cases (synucleinopathies, n = 3), comparing results from CSF and serum measurements from the same patients. CSF samples were measured on the previously published iRS surface, while serum samples were analyzed on a charge-modified sensor surface specifically designed for the serum matrix. Although the limited sample size does not allow meaningful statistical analysis, both body fluids reveal a general trend separating controls from αSyn misfolding cases. Median values indicate a separation between NPH without PD and PD/DLB of approximately 0.6 cm^-1^ for both CSF and serum, with serum CoM values consistently shifted to higher wavenumbers by about 1.0 cm^-1^. Box and whisker plots display the median (vertical line), interquartile range (boxes) and ±1 SD (whiskers). Reported values represent medians, with minima and maxima as follows: 1640.1–1641.3 (NPH; CSF), 1638.9–1639.9 (PD/DLB; CSF), 1641.3–1642.1 (NPH; serum), 1640.1–1640.7 (PD/DLB; serum). Abbreviations: αSyn alpha-synuclein, CoM center of mass, CSF cerebrospinal fluid, DLB dementia with Lewy bodies, iRS immuno-infrared-sensor, NPH normal pressure hydrocephalus (without PD), PD Parkinson’s disease, SD standard deviation

### Discovery and validation cohort

#### Study cohort description

In total, 127 blood serum samples from patients treated at the Paracelsus-Elena-Klinik Kassel, a large German movement disorder hospital, were analyzed using the iRS platform (*Tab. 1*). The samples consisted of a discovery cohort (n = 58) and an independent validation cohort (n = 69). In the discovery cohort, 18 individuals were diagnosed with synucleinopathies, namely PD (n = 9), MSA (n = 7), DLB (n = 1) or iRBD (n = 1) while 40 individuals served as controls, including healthy control (HC) participants (n = 28) and patients with other neurological disorders in which αSyn misfolding is not considered to be the pathophysiologic hallmark (NPH without PD, n = 11; essential tremor (ET), n = 1). The validation cohort comprised 56 individuals diagnosed with PD (n = 52), DLB (n = 3) or iRBD (n = 1) and 13 control individuals with neurological disorders not associated with αSyn misfolding (NPH without PD, n = 1; ET, n = 5; vascular PD (vaPD), n = 5; movement disorder without PD (noPD), n = 2). Diagnoses associated with αSyn misfolding (PD, MSA, DLB and iRBD) were classified as synucleinopathies, whereas all other diagnoses were considered misfolding negative and assigned to the disease control group. Clinical and demographic parameters, including age and sex, were available for all 127 individuals and statistical analyses were adjusted for age and sex. Motor symptom severity was assessed in 92 individuals (72%) using the Unified Parkinson’s Disease Rating Scale Part III (UPDRS-III). UPDRS-III assessments were not performed in 29 controls and 6 synucleinopathy cases. Smell testing was performed in 110 (87%) individuals across all diagnoses.

**Table. 1:** Demographic and clinical characteristics of study participants. The mean and SD for disease duration in the control group are based on individuals diagnosed with NPH without PD, ET, vaPD, or noPD, excluding the HC participants. Abbreviations: DLB dementia with Lewy Bodies, ET essential tremor, HC healthy control, iRBD isolated rapid eye movement sleep behavior disorder, MSA multiple system atrophy, noPD movement disorder without PD, NPH normal pressure hydrocephalus (without PD), PD Parkinson’s disease, SD standard deviation, UPDRS-III Unified Parkinson’s Disease Rating Scale Part III, vaPD vascular PD

| Parameter | Synucleinopathies |  |  | Controls |  |  |
| --- | --- | --- | --- | --- | --- | --- |
|  | Discovery | Validation | Combined | Discovery | Validation | Combined |
| **N total** | n = 18 | n = 56 | n = 74 | n = 40 | n = 13 | n = 53 |
| **Clinical Diagnosis** | PD = 9<br>MSA = 7<br>DLB = 1<br>iRBD = 1 | PD = 52<br>DLB = 3<br>iRBD = 1 | PD = 61<br>MSA = 7<br>DLB = 4<br>iRBD = 2 | HC = 28<br>NPH = 11<br>ET = 1 | NPH = 1<br>ET = 5<br>vaPD = 5<br>noPD = 2 | HC = 28<br>NPH = 12<br>ET = 6<br>vaPD = 5<br>noPD = 2 |
| **Age [years]**<br>(mean ± SD) | 69.7 ± 6.9 | 68.0 ± 9.7 | 68.4 ± 9.1 | 75.0 ± 6.7 | 73.1 ± 10.8 | 74.5 ± 8.0 |
| **Female [%]** | 61.1 | 41.1 | 45.9 | 35.0 | 30.8 | 34.0 |
| **Disease Duration [years]**<br>(mean ± SD) | 6.2 ± 4.8 | 2.2 ± 2.0 | 3.2 ± 3.4 | 6.6 ± 8.8<br>(0 N/A, n = 12) | 3.1 ± 3.3<br>(1 N/A, n = 12) | 4.9 ± 6.9<br>(1 N/A, n = 24) |
| **UPDRS-III**<br>(mean ± SD) | 35.7 ± 13.9<br>(3 N/A, n = 15) | 25.5 ± 13.8<br>(1 N/A, n = 55) | 27.7 ± 14.4<br>(4 N/A, n = 70) | 25.0 ± 10.5<br>(31 N/A, n = 9) | 19.9 ± 12.5<br>(0 N/A, n = 13) | 22.0 ± 12.0<br>(31 N/A, n = 22) |
| **Smell Identification Test**<br>(mean % ± SD) | 56.1 ± 13.8<br>(7 N/A, n = 11) | 53.0 ± 26.1<br>(1 N/A, n = 55) | 53.5 ± 24.5<br>(8 N/A, n = 66) | 71.5 ± 20.6<br>(7 N/A, n = 33) | 69.7 ± 25.7<br>(2 N/A, n = 11) | 71.1 ± 22.0<br>(9 N/A, n = 44) |

#### Group averages and difference spectrum

In individuals diagnosed with synucleinopathies, increased αSyn misfolding was expected, which is reflected by a loss of random coil and α-helical content and a corresponding increase in β-sheet structures. The iRS measurements yielded an average amide I CoM of 1641.4 cm^-1^ for the synucleinopathy group and 1642.7 cm^-1^ for controls. The amide I CoM was downshifted by 1.3 cm^-1^ in the synucleinopathy group, indicating a higher proportion of β-sheet-containing αSyn species. A similar shift was also observed in spiked Serodos^®^ controls (*Figure 1*). To further analyze the spectral differences, the mean control spectrum was subtracted from the mean misfolding spectrum (*Extended Data Fig. 4A*). The resulting difference spectrum showed a negative band at 1657 cm^-1^ and a positive band at 1635 cm^-1^ with comparable integrated intensities in the amide I region (1700–1600 cm^-1^). The negative band at 1657 cm^-1^ corresponds to a decrease in random coil and α-helical structures, while the positive band at 1635 cm^-1^ indicates an increase in β-sheet conformers, respectively. Additionally, a smaller but significant positive feature was visible at 1685 cm^-1^, which corresponds to antiparallel β-sheet structures^37^, suggesting the appearance of oligomeric species^38^. In contrast to fibrillar structures that mostly consist of parallel β-sheets, oligomers typically include antiparallel β-sheet structures^38^. Band shifts were also observed in the amide II region of the difference spectrum (1600–1500 cm^-1^). A negative band at 1590 cm^-1^ indicates the decrease in random coil/α-helical structures and a positive band at 1529 cm^-1^ shows the increase in parallel and antiparallel β-sheet structures^39^. Signal intensities corresponding to structural changes in the amide II region are smaller and less significant than in the amide I region. These findings demonstrate that the iRS platform can detect αSyn misfolding in blood serum of synucleinopathy patients without the need for complex, error-prone or time-consuming sample preparation.

#### Study results and statistics of distinct spectral features

The difference spectrum of group-level changes in the amide region revealed three distinctive spectral features, which were subsequently evaluated for their diagnostic value in distinguishing synucleinopathies from controls. These features captured structural transitions between random coil/α-helical and β-sheet conformations within the amide I (1654/1639 ratio) and amide II (1529/1555 ratio) regions, as well as additional variations in antiparallel β-sheet content (1660/1684 ratio). Individual statistical analyses showed that each spectral feature significantly distinguished between misfolding-positive samples and controls (*Extended Data Fig. 4B*). The strongest diagnostic performance was observed for the amide I band shift in the 1654/1639 ratio (P = 6.94 × 10^-8^), followed by the 1660/1684 ratio (P = 6.97 × 10^-9^) and the 1529/1555 ratio (P = 4.38 × 10^-5^). Statistical significance was assessed using Mann-Whitney U testing due to the non-normal distribution of the data (*Supplementary Fig. 1*). ROC AUC analyses showed values of 0.82 (95% confidence interval (CI), 0.75–0.90) for the 1660/1684 ratio, 0.83 (95% CI, 0.76–0.91) for 1654/1639 and 0.79 (95% CI, 0.71–0.87) for 1529/1555 (*Extended Data Fig. 4C*).

#### Study results of combined spectral features

Statistical analysis of the three spectral features revealed that the amide I band (1654/1639 ratio) and the 1660/1684 ratio reflecting group-specific changes in antiparallel β-sheet structures provided the highest diagnostic performance. The amide II ratio (1529/1555) also reached statistical significance, indicating added value when combined. In CSF sample analyses, only the amide I band ratio was used to distinguish synucleinopathies from healthy and neurological controls. In this study using serum samples, we observed that additional information on αSyn misfolding can be derived from changes in the antiparallel β-sheet region as well as from changes in the amide II region. To identify the optimal combination of spectral information, we next performed a combined analysis of the identified markers. A scaled combination of the 1660/1684 and 1529/1555 ratios with the 1654/1639 ratio yielded significantly improved results, with statistical significance increasing to P = 9.93 × 10^-12^ upon inclusion of additional spectral information from antiparallel β-sheets and the amide II region (*Fig. 3A*). The combined model achieved an enhanced AUC of 0.88 (95% CI, 0.82–0.94) across the full dataset of 127 study participants (*Fig. 3B*). In contrast, a logistic regression model using only age and sex, without the structural information from the iRS readout, yielded an AUC of 0.71 (*Supplementary Fig. 2*), demonstrating the added diagnostic value of αSyn as structural biomarker. Across cohorts, the discovery dataset achieved an AUC of 0.89, while the validation cohort reached 0.92 (*Fig. 3C*).

**Fig. 3:**
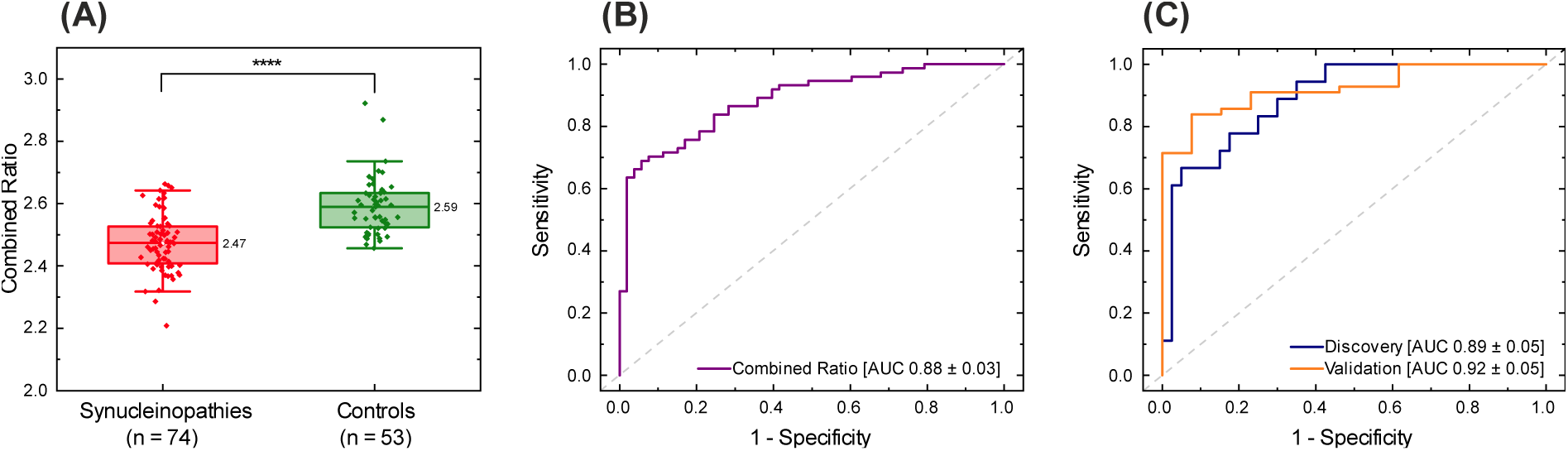
iRS analysis of αSyn secondary structure in serum using combined spectral ratios. (A) Boxplots comparing iRS-analyzed serum samples from individuals with synucleinopathies (n = 74) and controls (n = 53). A single threshold of 2.486 significantly distinguishes the two groups (Mann-Whitney U test, P = 9.93 × 10^-12^), based on a combined spectral ratio from Ratio2 (1654/1639 cm^-1^) + 0.3 × Ratio1 (1660/1684 cm^-1^) + 0.8 × Ratio3 (1529/1555 cm^-1^). Box and whisker plots show the median (vertical line), interquartile range (boxes) and ± 1 SD (whiskers). Each point represents one individual. Box minima and maxima range from 2.41–2.53 (synucleinopathies) and 2.52–2.63 (controls). (B) The ROC curve of the combined dataset (n = 127) shows an AUC of 0.88 ± 0.03 for differentiating synucleinopathies from controls. (C) ROC analyses of the discovery (n = 58) and validation (n = 69) cohorts yield AUCs of 0.89 ± 0.05 and 0.92 ± 0.05, respectively, confirming reproducible diagnostic performance. Abbreviations: αSyn alpha-synuclein, AUC area under the curve, iRS immuno-infrared-sensor, ROC receiver operating characteristic, SD standard deviation

#### Analyses of subgroup performance

To explore subgroup-level differences, we analyzed spectral ratios across all clinical subgroups (*Extended Data Fig. 5*). The control group (n = 53) consisted of different subgroups including HC, NPH without PD, ET, vaPD and noPD, all not associated with αSyn misfolding. The synucleinopathy group (n = 74) comprised individuals with PD (n = 61), MSA (n = 7), DLB (n = 4) and iRBD (n = 2), which are associated with αSyn misfolding. Boxplot analysis revealed substantial overlap within control and synucleinopathy subgroups, except for the two prodromal iRBD cases. Control subgroups showed median values of the combined ratio of 2.57 or higher (HC: 2.57, NPH: 2.57, ET: 2.60, vaPD: 2.64, noPD: 2.61), whereas misfolding subgroups showed lower median values (PD: 2.48, MSA: 2.49, DLB: 2.41, iRBD: 2.54). PD, MSA and DLB subgroups could be significantly distinguished from controls with P < 0.0001 for PD and DLB (PD: P = 1.62 × 10^-10^, DLB: P = 5.06 × 10^-6^) and P < 0.01 (P = 0.004) for MSA samples (*Extended Data Tab. 1*). In addition, no significant separation was observed between PD and DLB (P = 0.126) or PD and MSA (P = 0.526), whereas the comparison between DLB and MSA reached significance (P = 0.042), although small sample sizes (7 MSA, 4 DLB) limit statistical reliability.

#### Study results of a dual-threshold classification

To account for the overlap observed between misfolding-positive and -negative groups, we implemented a dual-threshold approach that categorized individuals into three groups (*Fig. 4*). This reflects the continuum of αSyn misfolding from healthy individuals to those with a final clinically confirmed synucleinopathy (PD, MSA, DLB), which a single cutoff value cannot fully represent. Therefore, participants with a combined ratio below 2.479 were classified as high misfolding (red), indicating a clear synucleinopathy profile, whereas those with ratios above 2.555 were classified as no misfolding (green), consistent with an unafflicted stage (*Fig. 4A*). Within the high misfolding group, 41 individuals were diagnosed with synucleinopathies, while two were false positive controls. In the no misfolding group, 33 individuals were controls and 11 were false negative. Individuals with ratios between the two thresholds (2.479–2.555) were assigned to an intermediate at-risk group with low misfolding (yellow), representing a transitional stage of emerging alpha Syn misfolding and may be on the path to clinical manifestation (n = 22 from synucleinopathy group and n = 18 from control group, 32% of all individuals). This low misfolding range reflects the gradual shift from healthy to disease-associated protein conformations. Individuals in this group may carry an elevated risk of developing synucleinopathies compared with those in the no misfolding group, although they do not yet exhibit a clear misfolding signature. When comparing only the high (n = 43, red) and no (n = 44, green) misfolding group, the difference was highly significant (P = 1.08 × 10^-10^), with an AUC of 0.94 (95% CI, 0.88–0.99), sensitivity of 88% and specificity of 89% (*Fig. 4B*).

**Fig. 4:**
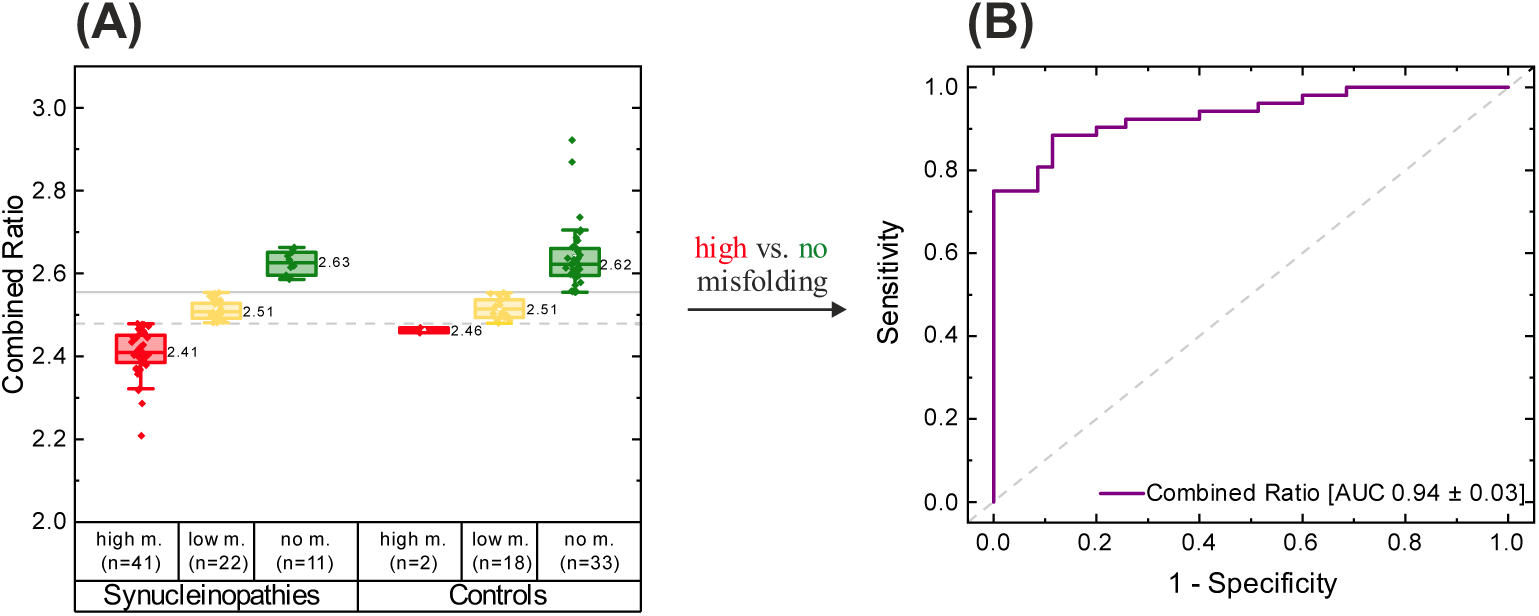
Dual-threshold classification of αSyn misfolding in serum using the iRS platform into high misfolding, no misfolding and an intermediate at-risk group with low misfolding. (A) The selected thresholds (2.479 and 2.555, grey lines) classify disease controls (controls; n = 53) and αSyn misfolding cases (synucleinopathies; n = 74) into three subgroups: high (red), low (inter; yellow) and no (green) misfolding. This scheme introduces an additional intermediate at-risk group to capture the overlap of αSyn misfolding pathology. Based on these thresholds, only 2 control individuals are assigned to the high misfolding group (false positives), while 18 fall into the intermediate range and 33 into the no misfolding group. Among synucleinopathy cases, 41 are classified as high misfolding, 22 as intermediate and 11 as no misfolding (false negatives). High and no misfolding groups could be significantly distinguished (Mann-Whitney U test, P = 1.08 × 10^-10^). Box and whisker plots show the median (vertical line), interquartile range (boxes) and ± 1 SD (whiskers). Each point represents one individual. Box minima and maxima range from 2.46–2.47 (controls, high misfolding), 2.49–2.54 (controls, low misfolding), 2.60–2.66 (controls, no misfolding), 2.39–2.45 (synucleinopathies, high misfolding), 2.49–2.53 (synucleinopathies, low misfolding) and 2.60–2.65 (synucleinopathies, no misfolding). (B) Based on clinical diagnoses, the ROC curve comparing high (n = 43) and no (n = 44) misfolding groups yields an AUC of 0.94 ± 0.03, with 88% sensitivity and 89% specificity. Abbreviations: αSyn alpha-synuclein, AUC area under the curve, iRS immuno-infrared-sensor, m. misfolding, ROC receiver operating characteristic, SD standard deviation

#### Diagnostic performance across early disease duration

To assess early diagnostic performance, we analyzed synucleinopathy samples collected within the first two years after symptom onset. Among these 44 cases, 35 (80%) were correctly classified as low or high misfolding using the previously defined thresholds (*Extended Data Fig. 6*). Focusing specifically on the first year, 22 out of 30 samples (73%) were correctly classified as low or high misfolding, while 8 (27%) were incorrectly classified as no misfolding. Notably, all synucleinopathy samples collected at the two-year mark, with one exception, were correctly assigned to the low or high misfolding group, indicating that the diagnostic performance of the iRS platform is already strong early in the disease course within very early clinical stages.

#### Combination of iRS readout with smell-function testing

Given the early onset of olfactory impairment in synucleinopathies, we assessed whether combining iRS readouts with smell-function testing improves diagnostic performance. Data from smell-function testing was available for 110 of 127 participants (87%), including 66 individuals with synucleinopathies and 44 controls as shown in detail in *Tab. 1*. Among the 17 participants without olfactory data, 8 were diagnosed with synucleinopathies and 9 were controls. Based on established smell-function cutoffs, >75% correct responses were classified as normosmia, 51–75% as hyposmia and ≤50% as anosmia. On average, synucleinopathy patients correctly identified 54% of odors, while control participants recognized 71%. A more detailed analysis and comparison of the iRS readout and smell-function testing, including absolute values and percentages for each group, are provided in *Extended Data Tab. 2*. Briefly, smell-function testing alone showed an age- and sex-adjusted AUC of 0.82 when samples were classified using the dual-threshold approach as described above (*Fig. 5A*). In comparison, the iRS achieved an AUC of 0.93 in the same subset, consistent with the performance of the full dataset (*Fig. 4*). Combining iRS readouts with smell function testing enhanced diagnostic performance, yielding an AUC of 0.96 (95% CI, 0.93–1.00), with 87% sensitivity and 97% specificity (*Fig. 5A*). When the iRS readout was plotted against smell-function testing, control samples predominantly showed low αSyn misfolding and preserved olfactory function (upper left quadrant), whereas synucleinopathy samples were mainly associated with high misfolding and hyposmia or anosmia and clustered in the lower right quadrant. (*Fig. 5B*). These results highlight the diagnostic strength of the iRS readout as a structure-based biomarker, with smell-function testing potentially offering complementary information and additional improvement.

**Fig. 5:**
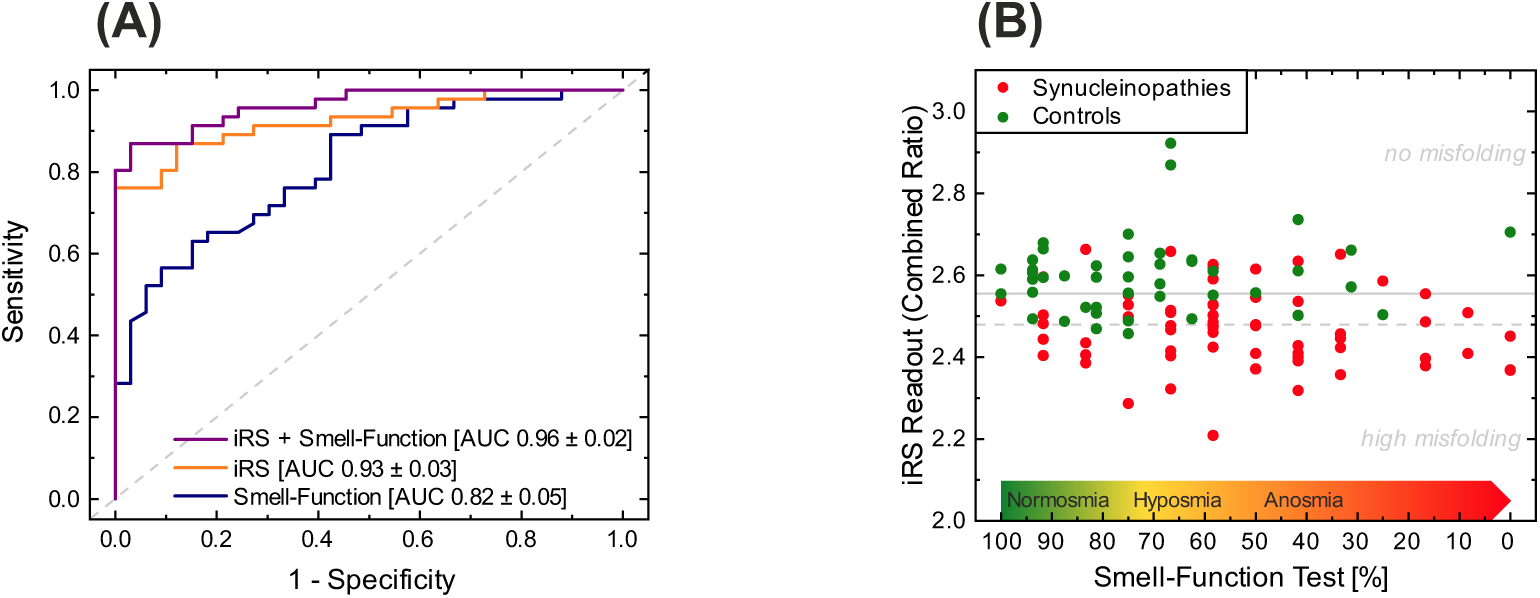
Diagnostic performance using combined iRS and smell-function test data. (A) ROC curve analysis demonstrates the diagnostic performance of the smell-function test (blue), the iRS readout (orange) and the combined approach (purple), with corresponding AUC values of 0.82 ± 0.05, 0.93 ± 0.03 and 0.96 ± 0.02, based on the 110 of 127 individuals with available smell-function test data. Samples were classified using dual-threshold cutoffs (2.479 and 2.555) with calculated AUC values of high misfolding and no misfolding. Combination of iRS readouts and smell-function testing, comprising 66 individuals with synucleinopathies (red dots) and 44 controls (green dots). Grey lines represent the iRS dual-threshold cutoffs (2.479 and 2.555), separating samples into high, low and no misfolding groups. Smell-function test results are classified based on the percentage of correctly identified odors: ≤ 50% correct is defined as anosmia, ≤ 75% correct as hyposmia. Abbreviations: AUC area under the curve, iRS immuno-infrared-sensor, ROC receiver operating characteristic

## Discussion

### Opening

Using the iRS platform, we show that misfolded αSyn can be directly detected in 250 µl of routinely collected serum, allowing the diagnosis of synucleinopathies without complex or error-prone sample preprocessing. Extending this technology from CSF to blood establishes αSyn misfolding as a promising high-performance structural biomarker in an easily accessible fluid for noninvasive diagnosis of synucleinopathies. Our findings provide direct evidence that disease-associated αSyn conformers are present in serum. In contrast to SAAs that yield a binary readout, the iRS platform resolves the progressive shift from α-helical/random-coil structures toward β-sheet-rich oligomers and fibrils. This structural progression reflects the continuum of αSyn misfolding from healthy to diseased states.

### Biological classification of PD and current αSyn biomarkers

The recently proposed SynNeurGe framework classifies PD based on pathological αSyn (S), neurodegeneration (N) and genetic background (G)^8^. Within this concept, misfolded αSyn as measured by SAA defines the “S⁺” state^8^. At the same time, another biological classification, the neuronal αSyn disease integrated staging system (NSD-ISS), has been proposed^7^. It extends the biological classification through a sequential staging framework that unifies PD and DLB under the shared concept of neuronal αSyn pathology^7^. This system has recently been applied in major PD cohorts, including the Parkinson’s Progression Markers Initiative (PPMI), PASADENA and SPARK^40^. Both frameworks highlight αSyn misfolding as the molecular hallmark of synucleinopathies and as a basis for biological classification. Currently, only CSF SAA positivity defines the S⁺ state in the NSD-ISS, whereas CSF and skin biopsies are recognized as validated matrices in the SynNeurGe framework^7,8^. However, skin biopsies have not been integrated into routine neurological practice and are more invasive than routinely applied blood sampling, while also showing reduced sensitivity in early PD^18–20^. Moreover, prodromal markers such as iRBD^41–43^ and olfactory loss^44,45^ help identify at-risk individuals who would benefit from additional noninvasive blood-based αSyn assays. Notably, Okuzumi et al. reported a serum-based SAA that distinguished individuals with PD and DLB from controls with high diagnostic accuracy, demonstrating the potential of peripheral αSyn as a blood-based biomarker^23^. Likewise, Kluge et al. developed a refined blood-based SAA that uses purified neuron-derived exosomes as αSyn seeds, achieving promising results not only in distinguishing PD from healthy individuals but also in monitoring disease progression and identifying early disease stages^24–27^. However, the robustness of these approaches and their comparability across laboratories remain under discussion, as no published blood-based study has yet achieved successful independent validation and reproducibility challenges have been noted by the community^14^.

### Challenges of blood-based αSyn assays

In general, SAAs require highly purified recombinant monomeric αSyn as substrate to achieve reproducible and standardized amplification results, highlighting the need for harmonized quality-control standards^12,14,46,47^. Even minimal blood contamination can disrupt assay performance in CSF. For example, Mammana et al. showed that contamination of ≥0.01% blood in CSF alters αSyn seeding kinetics^48^, underscoring the sensitivity of SAAs to matrix effects. Working with blood-based matrices is therefore particularly challenging, not only for SAAs but also for the iRS technique. The high protein concentration and complexity of plasma and serum can interfere with amplification efficiency^49,50^ or, in the case of the iRS platform, promote nonspecific protein adsorption on the biochip surface. Although total αSyn concentrations are higher in blood than in CSF^51^, the fraction of misfolded αSyn is substantially lower. As a result, pathological αSyn signals are diluted by abundant physiological αSyn and other interfering components^49,50,52^. Consequently, current blood-based SAAs depend heavily on preprocessing steps such as immunoprecipitation of αSyn^23^ or purification of neuron-derived exosomes^24–27^. Recently, silica nanoparticles were introduced to improve detection of misfolded αSyn in plasma without prior preprocessing by reducing inhibitory plasma effects on seeding activity^49^. While promising, these strategies still require further optimization and independent cohort validation before they can be implemented in routine diagnostics.

### Measuring αSyn misfolding using the iRS platform and contextualization

In contrast to SAAs, the iRS platform provides a label-free conformation-specific readout that directly measures the secondary structure distribution of immunocaptured αSyn using ATR-FTIR spectroscopy integrated within a microfluidic setup. Gao et al. reported an alternative label-free serum assay using surface plasmon resonance imaging (SPRi) with a peptoid ligand to distinguish PD cases from controls, achieving promising results in a small test cohort of 11 PD and 11 control individuals by quantifying total αSyn binding^53^. Using serum from routine blood draws and without requiring specialized preprocessing, the iRS platform resolved the secondary structure composition of αSyn and achieved an AUC of 0.88 (95% CI, 0.82–0.94) in distinguishing 74 synucleinopathy cases (PD, DLB, MSA and iRBD) from 53 controls (*Fig. 3*). Applying a dual-threshold classification between individuals with high (n = 43) and no (n = 44) misfolding further improved the diagnostic performance to an AUC of 0.94 (95% CI, 0.88–0.99), with 88% sensitivity and 89% specificity (*Fig. 4*). For comparison, Okuzumi et al. reported excellent diagnostic accuracies for PD and DLB, with AUCs of 0.96 and 0.90, respectively, whereas discrimination of MSA from controls was lower, with an AUC of 0.64, indicating restricted assay sensitivity for this form of synucleinopathy^23^. There is growing evidence that MSA-derived αSyn displays distinct filament folds and pathogenic mechanisms compared with PD and DLB, which likely contribute to differences in seed amplification efficiency and assay variability^54–56^. Subgroup analyses of iRS measurements yielded AUCs of 0.88 for PD, 0.97 for DLB and 0.91 for MSA (*Extended Data Tab. 1, Extended Data Fig. 5*). Although diagnostic accuracies for DLB and MSA are constrained by small sample sizes (DLB, n = 4; MSA, n = 7), the iRS platform shows potential to discriminate MSA samples from controls. Nonetheless, validation in an independent, MSA-focused study with larger cohorts will be essential. We believe that immunocapturing of αSyn is more comprehensive for detecting all conformers of αSyn than SAAs, as “seeding-competent” species are required for amplification, likely excluding early-stage disordered αSyn oligomers. Notably, these particular αSyn species, together with soluble protofibrils, are considered among the most neurotoxic^57^, emphasizing the need for reliable assay detection. There is spectroscopic evidence that we resolve oligomeric αSyn species, as the difference spectrum of group-level alterations in the amide region revealed an increase in antiparallel β-sheet structure within the synucleinopathy group. Using this spectral feature alone for performance evaluation yielded strong differentiation between synucleinopathies and controls, with an AUC of 0.82 (*Extended Data Fig. 4*), highlighting the contribution of oligomeric αSyn species in synuclein aggregation disorders. Recent post-mortem analyses have indicated that several leucine-rich repeat kinase 2 (LRRK2) mutation carriers lack classical Lewy body fibrillar αSyn inclusions but instead contain abundant particulate αSyn species, likely oligomeric^58,59^. Applying the iRS technology to LRRK2 individuals could help determine whether they exhibit an increased oligomeric burden, as both oligomeric and fibrillar αSyn species can be detected by the iRS assay. Moreover, our findings demonstrate that the diagnostic performance of the iRS platform is robust in early disease stages, with detectable αSyn misfolding observed in the majority of synucleinopathy cases within a few months of motor symptom onset (*Extended Data Fig. 6*). Because the iRS assay may detect molecular changes even before symptom onset, it is possible that some individuals classified as false negative in the clinical dataset are not truly unaffected but are instead in a prodromal stage that has not yet reached clinical diagnosability. Additionally, combining the iRS readout with accessible and cost-effective olfactory testing further enhanced diagnostic accuracy to an AUC of 0.96 (*Fig 5*). This two-step approach, combining simple olfactory and blood testing, enables early, noninvasive detection of synucleinopathies, democratizes access to diagnosis for individuals at elevated risk and supports large-scale population screening efforts currently underway in preparation for prevention trials. It offers a powerful biological classification of synucleinopathies and facilitates the very early identification of these individuals. This strategy will support earlier, more precise clinical intervention, offering a significant advantage over the symptom-based diagnosis currently used.

### Discussion of controls and methodology

Previous iRS measurements demonstrated that Aβ misfolding in both CSF and plasma serves as a highly accurate structural biomarker for the early and differential classification of AD^29–34^. Building on this foundation, we extended the iRS platform technology from AD to PD. For the first time, we showed the conversion of αSyn from random-coil/α-helical to β-sheet-enriched conformers in unmodified CSF^35^ and now confirmed this finding also in blood. To transfer the iRS-detected αSyn conversion from CSF to a minimally invasive, blood-based assay, we characterized the surface performance and αSyn target specificity (*Fig. 1, Extended Data Fig. 2/3*). Repeated measurements demonstrated robust reproducibility of the amide I CoM, with a standard deviation of 0.27 cm^-1^ (*Supplementary Fig. 3*). With the improved functionalized surface, we first analyzed paired CSF and serum samples. Both matrices showed clear group separation between PD/DLB and NPH controls in a small test cohort (*Fig. 2*). To further improve measurement precision, we are currently transferring the workflow from home-built FTIR-based instruments to commercial iRS systems (betaSENSE GmbH). These systems utilize Quantum Cascade IR-laser (QCL) technology, which provides more precise infrared measurements than FTIR and enables platform upscaling through parallel measurements due to the high-intensity laser source. In addition, the improved microfluidics in the betaSENSE iRS instruments show superior temperature stability, with significantly reduced baseline drift and more precise sample flow. The improvement in laser measurement and the microfluidic system will ultimately enhance accuracy and robustness. The iRS-QCL-based instruments are CE-certified and clinical study measurements under GCLP (Good Clinical Laboratory Practice) conditions are possible.

### Summary and outlook

Our findings highlight the potential of the iRS platform as a powerful and versatile tool for detecting disease-associated αSyn misfolding directly in small volumes of routinely collected blood serum. By extending this technology from CSF to blood serum, we demonstrate that αSyn misfolding can serve as a robust, noninvasive biomarker across the entire spectrum of synucleinopathies. The structure-specific readout of the iRS platform provides sensitive stage-specific insights into the disease continuum, going beyond the binary results obtained from SAAs. With additional clinical validation in larger, genetically defined cohorts, including individuals at earlier NSD-ISS stage 2 who are at risk of conversion, the iRS platform has the potential to become a broadly applicable tool for precise risk stratification and disease progression monitoring in synucleinopathies. This includes well-characterized cohorts such as the PPMI, where CSF SAA and dopamine transporter imaging results are available. Ultimately, this will support the enrollment of participants in future prevention trials and the development of therapeutic drugs, like approaches used for AD.

## Methods

### Materials, technical equipment and software

Antibodies, assay materials and components, kits, equipment and software used in this study are listed in *Tab. 2*. AssayDefender^®^ (CANDOR Bioscience GmbH, Wangen, Germany) is a commercially available blocking solution used to suppress nonspecific binding in immunoassays. Serum samples were diluted 1:1 with AssayDefender^®^ to reduce matrix-related interference during iRS measurements. Serodos^®^ (HUMAN Gesellschaft für Biochemica und Diagnostica mbH, Wiesbaden, Germany) is a certified, lyophilized human serum reference material with analyte concentrations within the normal physiological range. It serves as a quality control or calibration standard for clinical chemistry and immunoassay applications. The material is reconstituted with distilled water prior to use and provides a well-characterized serum matrix containing endogenous proteins, lipids and metabolites representative of native human serum.

**Table. 2.** Materials, reagents, equipment and software.

| Reagent | Source | Identifier or catalog number |
| --- | --- | --- |
| **Antibodies** |  |  |
| Capture antibody fragment: scFv, epitope aa 131-140 (iRS analysis) | betaSENSE GmbH | N/A |
| **Assay materials and components** |  |  |
| αSyn monomers, oligomers, PFFs | StressMarq Biosciences, Inc. | Cat # 321, Cat # 466, Cat # 322 |
| Albumin from human serum | Sigma-Aldrich | Article No.: A1653 |
| AssayDefender® | CANDOR Bioscience GmbH | Article No.: 180 |
| Serodos® | HUMAN Gesellschaft für Biochemica und Diagnostica mbH | REF: 13951 |
| **Kits** |  |  |
| LEGEND MAX™ Human αSyn (Colorimetric) ELISA Kit | BioLegend, Inc. | Cat # 448607 |
| Sniffin' Sticks test kit | Burghart Messtechnik GmbH | N/A |
| **Equipment** |  |  |
| Vertex 80 V | Bruker Optics | N/A |
| iRS flow system | home-built | N/A |
| Microplate shaker with lid | VWR International, LLC | N/A |
| CLARIOstar® Plus | BMG LABTECH GmbH | N/A |
| **Software** |  |  |
| MARS Data Analysis | BMG LABTECH GmbH | N/A |
| MATLAB R2019a | The MathWorks, Inc. | N/A |
| OriginPro 2019 | OriginLab Corporation | N/A |
| OPUS 7.2 | Bruker | N/A |
| AVASYS | in-house software | N/A |
| FileViewer AVASYS | in-house software | N/A |

### Study cohorts and sample collection

All participants provided written informed consent and are part of the De Novo Parkinson (DeNoPa) study or the Kassel cohort study. Both studies were approved by local ethics committee, respectively (approval no. FF89/2008 and FF38/2016). Recruitment, study design and sample collection have been published previously^60–63^. Serum was collected by venous puncture under fasting conditions in the morning and centrifuged for 10 min at 20° C and 2500 × g. Serum was aliquoted and stored at -80° C within 30 min after collection. *Tab. 1* presents the study cohorts along with demographic (age, sex) and clinical characteristics, including disease duration and additional clinical measures (UPDRS-III scores and smell test results), where available. The assessment of olfactory function was performed using the commercial *Sniffin’ Sticks* test kit (Burghart Messtechnik GmbH, Holm, Germany) according to the manufacturer’s guidelines. Participants were presented with either 12 or 16 distinct odors and asked to identify each odor from four multiple-choice options. Olfactory performance was classified based on the proportion of correctly identified odors: >75% indicating normosmia, 51–75% hyposmia and ≤50% anosmia. After data acquisition and analysis, results were shared with collaborating sites, which provided the corresponding clinical information for performance evaluation.

### Determination of αSyn-misfolding using the iRS platform

The misfolding of αSyn from blood serum samples was performed via the iRS platform technology. As a readout, the structure-sensitive amide I absorbance band was analyzed, which primarily arises from the C=O stretching vibration of the peptide backbone. In addition, the amide II band was monitored that mostly results from the N–H bending vibration of peptide bonds in the protein backbone and serves as an indicator for the protein amount. Variations in secondary structure induce shifts in the amide I band. While random coil and α-helical structures shift the maximum of the amide I band to higher wavenumbers (around 1650 cm^-1^), β-sheet-rich conformations induce a shift to lower wavenumbers (around 1620 cm^-1^). As a result, differences in the secondary distribution of proteins, such as the examined biomarker can be assessed. The ATR technique enables measurements in aqueous environments by limiting the penetration depth to approximately 500 nm, thereby reducing the background absorbance from water molecules, as the O–H bending vibration is several orders of magnitude larger. An ATR unit was integrated into a Vertex 80 V FTIR spectrometer (Bruker Optics, Ettlingen, Germany) equipped with a mercury cadmium telluride (MCT) detector and a middle infrared (MIR) source. Measurement parameters were used as described previously and sample measurements were performed in a multichannel flow-through setup^29,30^. Spectra were analyzed by in-house software together with Matlab and Python scripts.

### Functionalization of the iRS surface

The previously established iRS surface functionalization, as described in Schuler et al.^35^ and Gerwert et al.^64^ and in patents WO2024003213A1 and WO2024003214A3, was refined to ensure stability and specificity under the challenging conditions of blood serum, including high serum protein concentrations and other interfering components. A detailed description of this newly developed serum-optimized surface is protected by a pending patent application and will become publicly accessible upon patent publication. Briefly, the surface was specifically modified in terms of surface charge and inertness using specific polyethylene glycol (PEG)-containing linker molecules to further reduce nonspecific binding, while ensuring efficient target binding. Surface functionalization was performed using both dipping and flow-through setups. For serum measurements, surfaces were freshly prepared the day before each experiment. Quality control was ensured by the iRS setup, verifying each surface component through its characteristic absorbance bands. Immobilization of antibody fragments was conducted directly before the sample measurement. Background spectra were recorded after stabilization in PBS buffer and blood serum samples (250 µl) were applied in a 1:1 dilution with AssayDefender™ (CANDOR Bioscience GmbH, Wangen, Germany). Samples were circulated for one hour over the surface, followed by a subsequent wash for two hours with PBS buffer. Repeated spectra were recorded during every measurement step (20 s/spectrum) with background spectra taken before every new step (60 s/spectrum). Averaged wash spectra (spectra 2-18) were taken for evaluation.

### Antibody production, labeling and immobilization

The recombinant scFv constructs targeting the αSyn epitope at amino acids 131–140 (sequence from betaSENSE) were expressed using the ExpiCHO™ Expression System (ThermoFisher Scientific Inc.), following instruction of the manufacturer. The sequence was cloned into the pcDNA™ 3.4 TOPO™ expression vector and transfected into ExpiCHO-S™ cells (ThermoFisher Scientific Inc.), which were cultured in a high-yield-optimized, serum-free and chemically defined medium. To enable site-specific binding to the functionalized surface, a C-terminal cysteine was added to the constructs, allowing thiol-maleimide conjugation with EZ-Link™ Maleimid-PEG4-DBCO (ThermoFisher Scientific Inc.). Conjugated scFv was evaluated using ELISA and fluorescence polarization to confirm binding capability to αSyn monomers, oligomers and fibrils. Until further use, the conjugated scFv was aliquoted, stored in PBS at -80 °C and thawed freshly before each sample measurement.

### αSyn quantification in serum using ELISA

αSyn amounts in original blood serum samples and measurement supernatants were quantified using the LEGEND MAX™ Human αSyn (Colorimetric) ELISA Kit (BioLegend Inc., San Diego, US). All procedures followed the manufacturer’s instructions, with a final dilution of 1:100 applied to serum samples. For the measurement supernatants, the predilution caused by the flow system volume was accounted in the calculations. All blood serum original samples, measurement supernatants, standard curves and ELISA controls were measured in at least duplicates. Washing steps were performed using the plate washer Wellwash™ Versa and absorbance at 450 nm (with background at 570 nm) was recorded with the CLARIOstar Plus (BMG LABTECH GmbH, Ortenburg, Germany). The plate reader was configured to 22 flashes per well per cycle, with double orbital shaking at 300 rpm for 5 s before reading. Data analysis was performed with the included MARS software and involved blank correction, background subtraction and a 4-parameter logistic fit of the standard curves. The standard curves were then used to calculate αSyn amounts in both blood serum original samples and measurement supernatants.

### Spectra processing, data analysis and statistical evaluation

Spectra processing and data analysis were performed with an in-house developed software AVASYS, along with MATLAB and Python scripts. For evaluation and quality control, both kinetics and single absorbance band positions (e.g., 1550 cm^-1^ for amide II, representing protein amount), as well as whole absorbance spectra (4000–1450 cm^-1^), were considered. Spectral quality was assessed using the signal-to-noise (S/N) ratio, the amide I/amide II ratio (indicator of potential water absorbance variability) and the spectral baseline offset at 3900 cm^-1^. Evaluation spectra were generated by averaging spectra 2–18 of the sample wash step and the corresponding difference spectrum was obtained by recalculating with the average of 25 background spectra. Subsequent corrections included scaled subtraction of a water vapor spectrum and linear baseline correction with fixed points at 2700 and 1855 cm^-1^. Spectra were smoothed using Gaussian smoothing with a smoothing range of 7 cm^-1^. To determine spectral ratios, evaluation spectra of control and synucleinopathy groups were averaged and area-normalized across the amide I and II regions. Combined ratios were always calculated through scaled addition of single ratios. All smoothing, normalization and ratio calculations were performed using self-developed Python scripts. Initial spectral analyses, including comparisons between CSF and serum samples, were performed using the amide I center of mass (CoM), which was calculated from the upper 10% and 20% of the 1700–1600 cm^-1^ region and then averaged. The CoM was chosen for evaluation because it is more stable and robust against artefacts and water vapor interference than the absolute band maximum. Statistical analyses were conducted in OriginPro 2019. Group separation was evaluated using ROC analysis with AUC calculation and statistical significance was assessed using Mann-Whitney U testing (α = 0.05). For ROC AUC estimation, foregoing logistic regression was performed in Python with the clinical diagnosis as dependent variable and age, sex and the iRS readout as independent variables. Additional models included only age and sex or combination of age, sex, smell-function test results and iRS readouts. The probabilities received from these models were further used for ROC AUC calculations.

## Data Availability

All data produced in the present study are available upon reasonable request to the authors

## Acknowledgements

The authors thank Prof. Dr. Ralf Gold for his continuous support, including the provision of well-characterized samples and expert advice as an experienced neurologist. This research was funded by the Center for Protein Diagnostics (ProDi) and the Ministry of Culture and Science of North Rhine-Westphalia.

## Author contributions

Lennart Langenhoff, Jonas Simon and Sandrina Weber are shared first authors.

<u>Lennart Langenhoff:</u> Conceptualization; Methodology; Formal analysis; Investigation; Data curation; Writing – Original Draft; Writing – Review & Editing.

<u>Jonas Simon:</u> Conceptualization; Methodology; Formal analysis; Investigation; Data curation; Visualization; Writing – Original Draft; Writing – Review & Editing.

<u>Sandrina Weber:</u> Investigation; Resources; Data curation; Formal analysis; Writing – Review & Editing.

<u>Diana Hubert:</u> Methodology; Investigation; Resources

<u>Martin Schuler:</u> Methodology; Formal analysis; Investigation

<u>Marvin Mann:</u> Methodology; Investigation

<u>Vuk Puzovic:</u> Investigation.

<u>Grischa Gerwert:</u> Methodology; Investigation

<u>Adrian Höveler:</u> Methodology; Investigation

<u>Léon Beyer:</u> Methodology; Investigation

<u>Lars Tönges:</u> Supervision

<u>Carsten Kötting:</u> Methodology; Supervision; Writing – Review & Editing.

<u>Jörn Güldenhaupt:</u> Methodology; Validation; Formal analysis; Investigation; Data curation; Writing – Review & Editing.

<u>Brit Mollenhauer:</u> Conceptualization; Resources; Supervision; Funding acquisition; Data curation; Formal analysis; Writing – Review & Editing.

<u>Klaus Gerwert:</u> Conceptualization; Resources; Supervision; Project administration; Funding acquisition; Writing – Original Draft; Writing – Review & Editing.

## Additional information

Extended data

Supplementary information Disclaimer

This manuscript is a preprint and has not undergone peer review. The findings should not be interpreted as clinical guidance.

---

**Extended Data Fig. 1:**
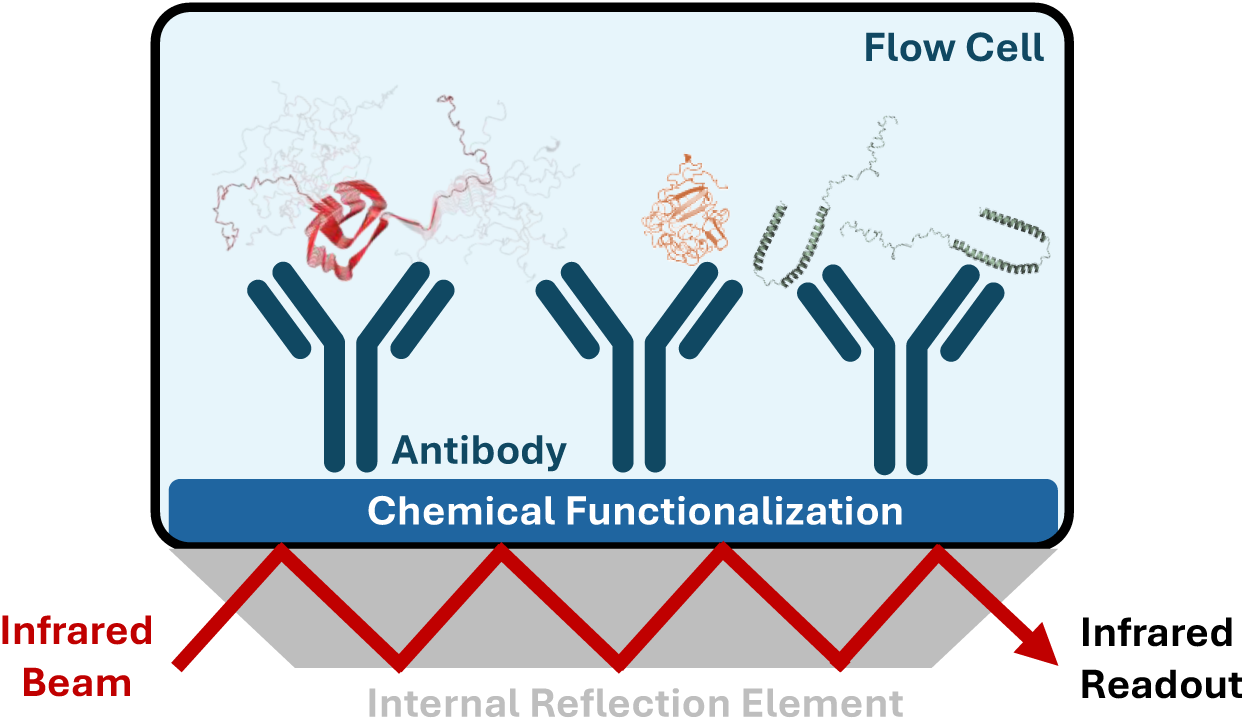
Schematic representation of the iRS biochip. All αSyn conformers (monomers, oligomers, fibrils) are extracted from the body fluid with specific capture antibodies on the iRS surface. Chemical linker compounds prevent unspecific binding to the surface. An infrared beam monitors the absorbance of all components bound to the ATR surface. To reveal only the absorbance of αSyn, previously measured background spectra of the functionalized surface are subtracted. The readout is the structure sensitive amide I band indicating the secondary structure distribution of the bound biomarker. Additional information can be received from the amide II band, which indicates the amount of the bound biomarker. Abbreviations: αSyn alpha-synuclein, ATR attenuated total reflection, iRS immuno-infrared-sensor

**Extended Data Fig. 2:**
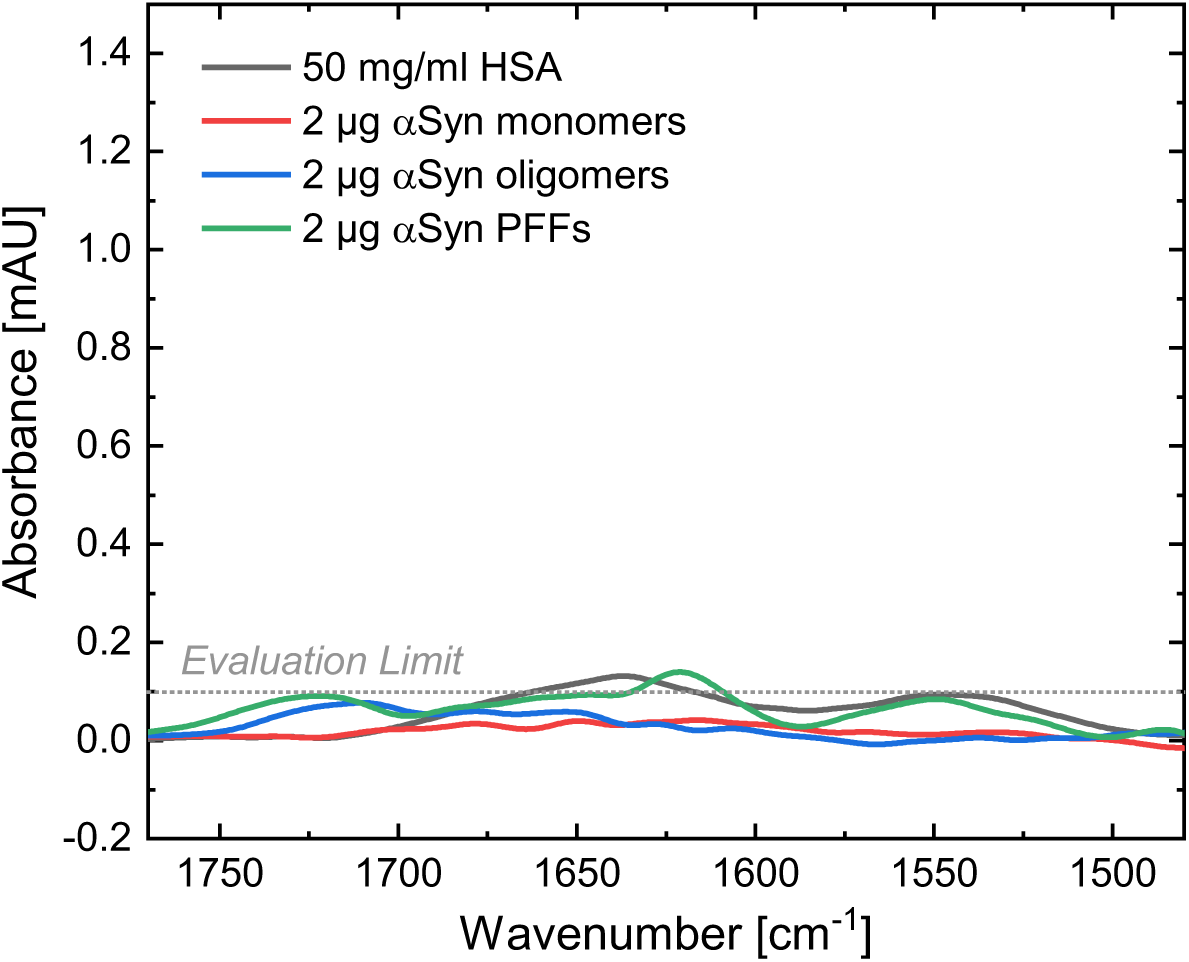
Characterization of nonspecific adsorption of αSyn conformers and HSA at high concentrations on a charge-modified iRS surface for serum measurements. The optimized iRS biochip without capture antibodies demonstrates high surface performance, as no signals are detected when exposed to large amounts (2 µg) of αSyn conformers (monomers, oligomers, PFFs) or to a high concentration (50 mg/ml) of HSA, the most abundant serum protein. Only minimal nonspecific adsorption is observed for PFFs and HSA, with amide II values remaining below the evaluation limit defined by the S/N ratio. These findings confirm sufficient surface resistance to nonspecific binding, as physiological protein concentrations are below tested levels. Abbreviations: αSyn alpha-synuclein, HSA human serum albumin, iRS immuno-infrared-sensor, PFFs preformed fibrils, S/N signal-to-noise

**Extended Data Fig. 3:**
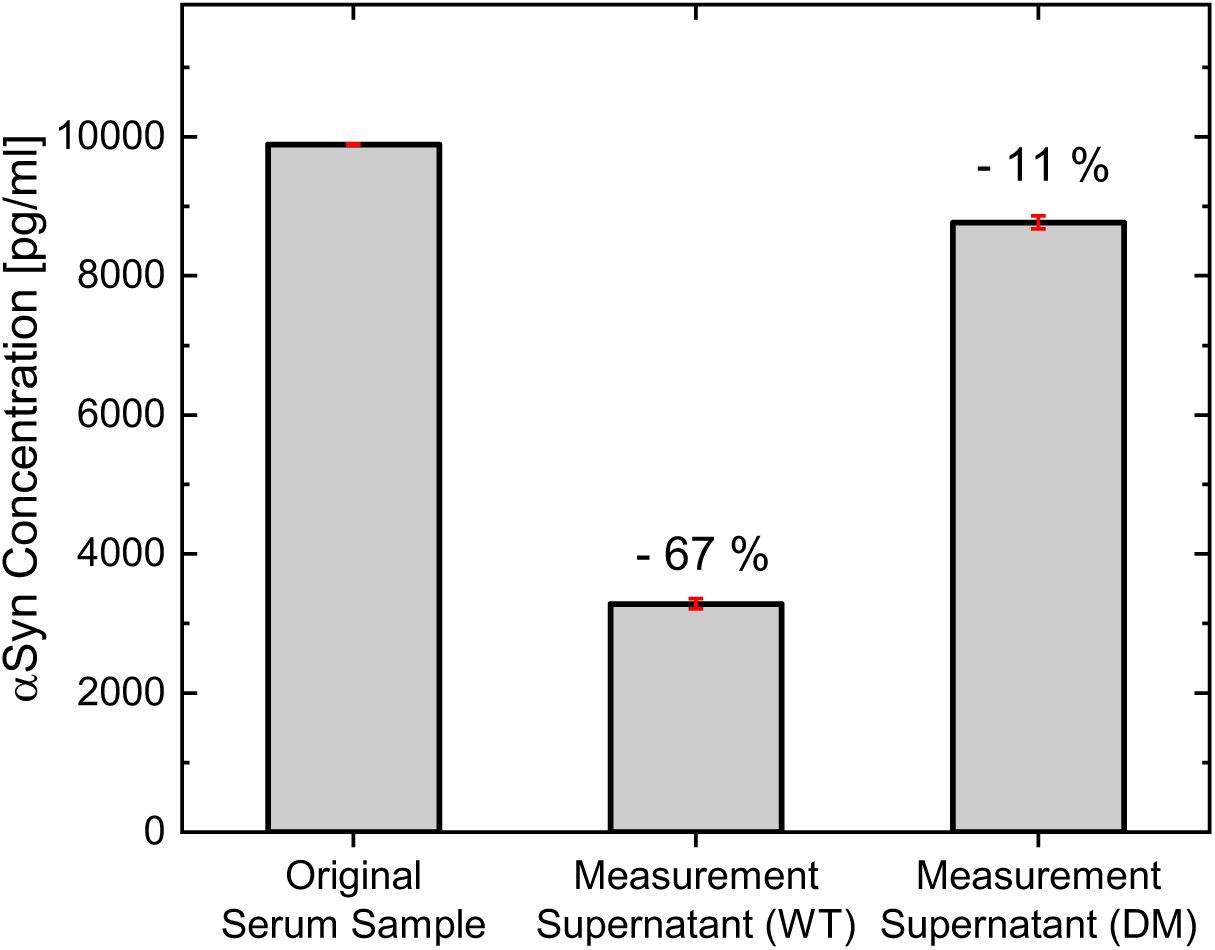
Indirect ELISA characterization of the iRS platform for quantifying surface-bound αSyn. Quantification of αSyn concentrations before and after measuring one exemplary serum sample on the iRS using surfaces functionalized with either an active antibody wildtype (WT) fragment or a binding-deficient dead mutant (DM) as a negative control. The original serum sample contains 9.892 ng/mL of αSyn. Following iRS measurement, supernatants contain 3.283 ng/mL (WT) and 8.767 ng/mL (DM), indicating approximately 67% specific binding to the WT-functionalized surface. The 11% reduction on the DM-functionalized surface reflects nonspecific losses within the system volume. Samples and supernatants from the ATR were diluted to fit the standard range and measured in ELISA duplicates. Error bars (red) represent the standard deviation of duplicates and gray columns indicate the mean values. Abbreviations: αSyn alpha-synuclein, DM dead mutant, ELISA enzyme-linked immunosorbent assay, iRS immuno-infrared-sensor, WT wildtype

**Extended Data Fig. 4:**
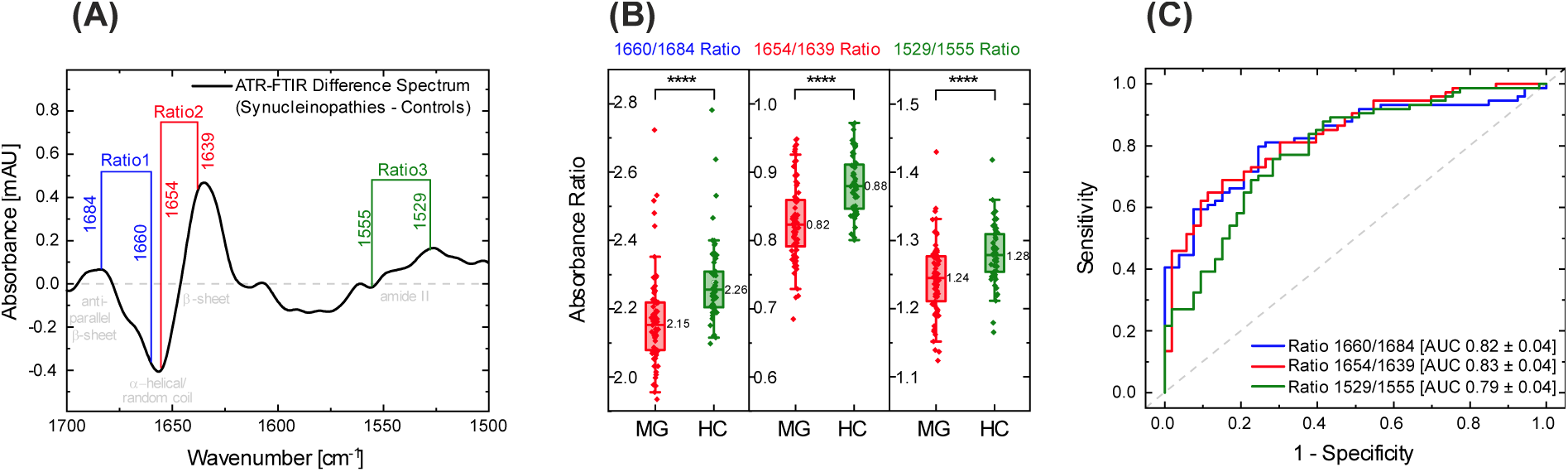
Analysis of αSyn secondary structure distribution in serum using the iRS platform. (A) The difference spectrum of group-level alterations in the amide region was obtained by subtracting averaged and normalized spectra of the misfolding group (n = 74) from those of the control group (n = 53). The resulting spectrum shows negative absorbance values at 1657 cm^-1^ and between 1605 and 1552 cm^-1^ and positive absorbance values at 1684 cm^-1^, 1635 cm^-1^ and 1529 cm^-1^, indicating a decrease in α-helical/random-coil structures and an increase in β-sheet structures in the PD/MSA/DLB and iRBD samples compared with controls. Transitions from positive to negative absorbance were considered as distinct spectral features of misfolding-related changes and three ratios (Ratio1: 1660/1684, Ratio2: 1654/1639, Ratio3: 1529/1555) were identified. The ratio of these signals was subsequently used to distinguish misfolding cases from controls. (B) Boxplots of the three spectral ratios, each derived from distinct spectral features, are shown for the combined dataset of 127 individuals, including 74 synucleinopathies (red) and 53 controls (green), with each point representing one participant. A single individual threshold was determined for each ratio: 2.186 for 1660/1684, 0.832 for 1654/1639 and 1.252 for 1529/1555. Mann-Whitney U testing confirmed statistically significant group differences (1660/1684: P = 6.97 × 10^-9^; 1654/1639: P = 6.94 × 10^-8^; 1529/1555: P = 4.38 × 10^-5^). Box-and-whisker plots indicate the median (vertical line), interquartile range (boxes) and ± 1 SD (whiskers). For the 1660/1684, 1654/1639 and 1529/1555 ratios, box minima and maxima in the misfolding group range from 2.08–2.22, 0.79–0.86 and 1.21–1.28, respectively, with control group values ranging from 2.20–2.31, 0.85–0.91 and 1.25–1.31. (C) ROC curves for the combined dataset (n = 127), based on the three spectral ratios, yield AUCs of 0.82 ± 0.04, 0.83 ± 0.04 and 0.79 ± 0.04 for classifying misfolding cases (synucleinopathies) versus controls using the 1660/1684, 1654/1639 and 1529/1555 ratios, respectively. Abbreviations: αSyn alpha-synuclein, AUC area under the curve, DLB dementia with Lewy bodies, HC healthy control, iRBD isolated rapid eye movement sleep behavior disorder, iRS immuno-infrared-sensor, MG misfolding group, MSA multiple system atrophy, PD Parkinson’s disease, ROC receiver operating characteristic, SD standard deviation

**Extended Data Fig. 5:**
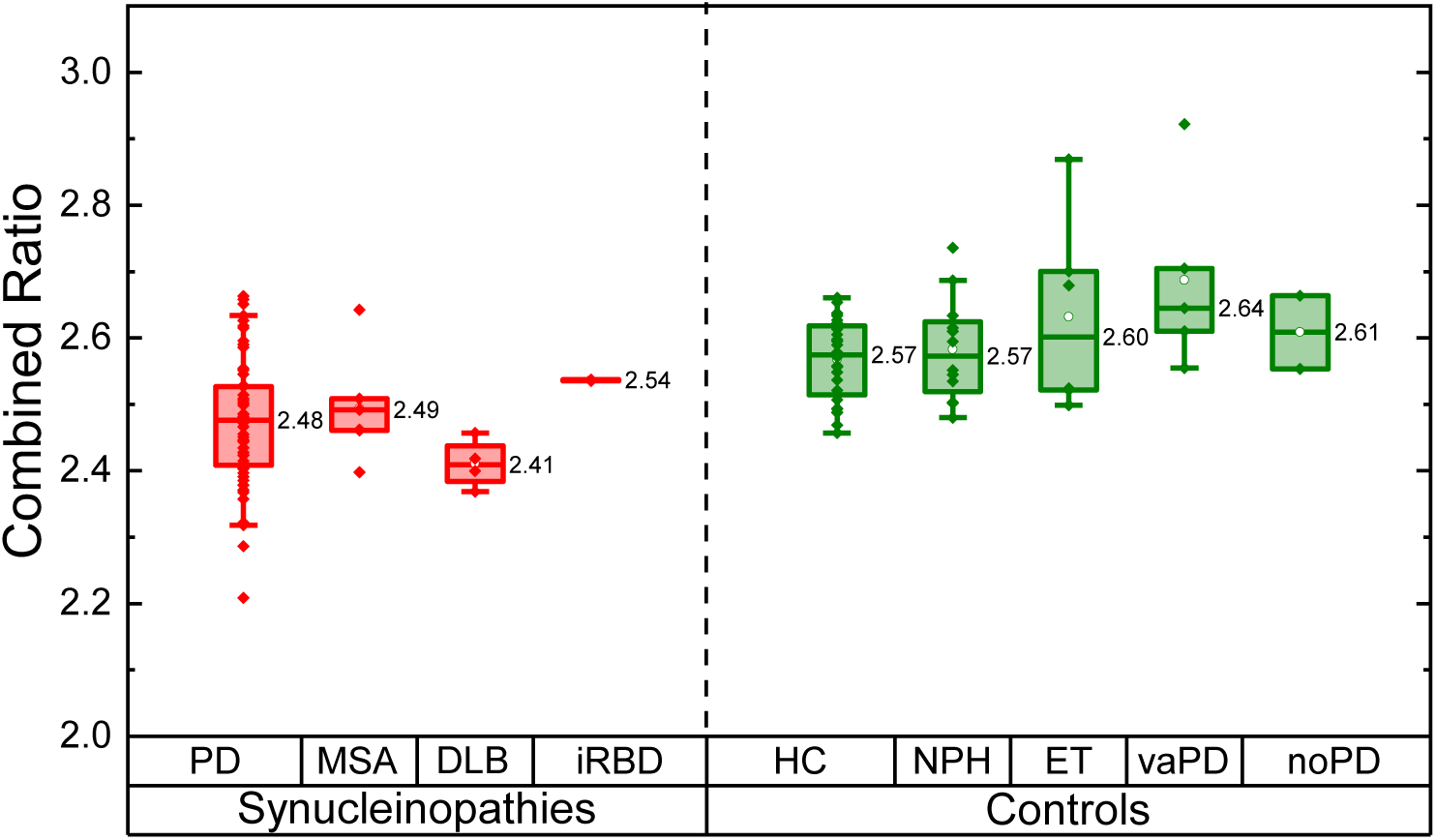
**Diagnostic subgroup comparison of controls and synucleinopathies using the iRS platform**. Synucleinopathies (n = 74) include PD (n = 61), MSA (n = 7), DLB (n = 4) and iRBD (n = 2). The control group (n = 53) comprises various diagnoses, including HC (n = 28), NPH without PD (n = 12), ET (n = 6), vaPD (n = 5) and noPD (n = 2), in which αSyn misfolding is considered highly unlikely. Within both control and synucleinopathy subgroups, boxplots show substantial overlap in values, particularly at the median, except for the two iRBD cases. Apart from the HC and PD groups, sample sizes were too small to support meaningful statistical comparisons. Nonetheless, DLB samples tended to show lower values than MSA samples and the PD group median. Box and whisker plots display the median (vertical line), interquartile range (boxes) and ± 1 SD (whiskers). Each point represents one individual. Reported values indicate subgroup medians, with box minima and maxima as follows: 2.41–2.53 (PD), 2.46–2.51 (MSA), 2.38–2.44 (DLB), 2.52–2.62 (NPH), 2.51–2.62 (HC), 2.52–2.70 (ET), 2.61–2.70 (vaPD) and 2.55–2.66 (noPD). Abbreviations: αSyn alpha-synuclein, DLB dementia with Lewy bodies, ET essential tremor, HC healthy control, iRBD isolated rapid eye movement sleep behavior disorder, iRS immuno-infrared-sensor, MSA multiple system atrophy, noPD movement disorder without PD, NPH normal pressure hydrocephalus (without PD), PD Parkinson’s disease, SD standard deviation, vaPD vascular Parkinson

**Extended Data Fig. 6:**
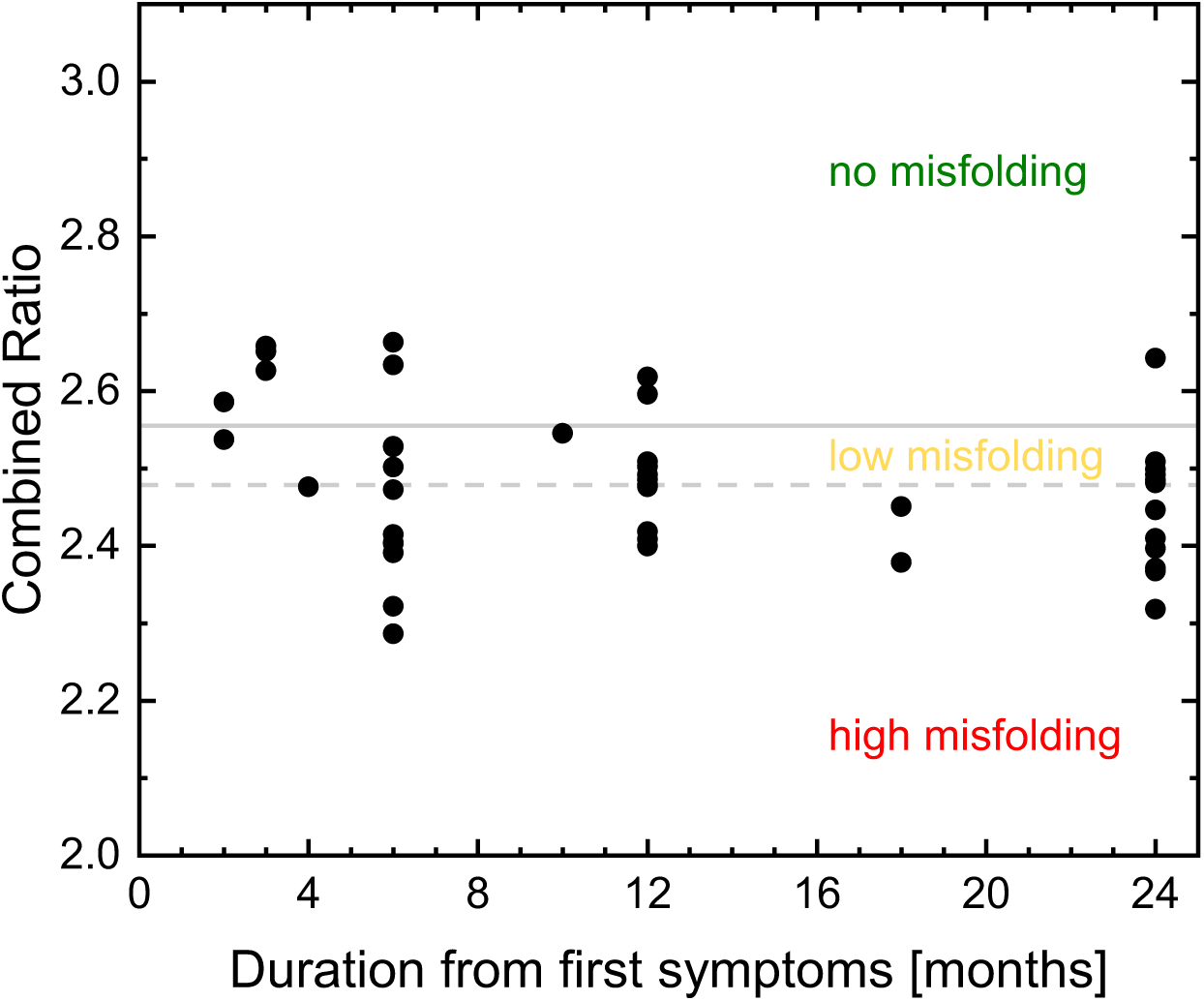
**Early classification of synucleinopathies using the iRS platform within the first two years after symptom onset**. Classification of synucleinopathy serum samples using the iRS readout within the first two years after symptom onset. Among 44 early-stage cases with two years disease duration or less, 80% are assigned to the low or high misfolding group. Within the first year, 22 out of 30 cases (73%) show elevated misfolding. All but one sample collected at the two-year mark fall into the low or high misfolding group, highlighting the strong early diagnostic performance of the iRS technology, which further improves with longer disease duration. Grey lines indicate the dual threshold separation (2.479 and 2.555). Abbreviations: αSyn alpha-synuclein, iRS immuno-infrared-sensor

**Extended Data Tab. 1:** Statistical comparison between synucleinopathy subtypes and controls. αSyn misfolding readouts from synucleinopathies (PD, n = 61; DLB, n = 4; MSA, n = 7) are compared to the combined control group (n = 53) and with each other. The table presents ROC AUC values with standard errors (top rows) and statistical significance assessed using Mann-Whitney U testing (italicized values in lower rows). Abbreviations: αSyn alpha-synuclein, DLB dementia with Lewy bodies, MSA multiple system atrophy, NS not significant, PD Parkinson’s disease, ROC receiver operating characteristic area under the curve

|  | PD<br>(n = 61) | DLB<br>(n = 4) | MSA<br>(n = 7) | Controls<br>(n = 53) |
| --- | --- | --- | --- | --- |
| PD<br>(n = 61) | - | 0.52 ± 0.22<br>(NS)<br>0.126 | 0.56 ± 0.15<br>(NS)<br>0.526 | 0.88 ± 0.03<br>(****)<br>$1.62 \times 10^{-10}$ |
| DLB<br>(n = 4) | 0.52 ± 0.22<br>(NS)<br>0.126 | - | 0.50 ± 0.21<br>(*)<br>0.042 | 0.97 ± 0.07<br>(****)<br>$5.06 \times 10^{-6}$ |
| MSA<br>(n = 7) | 0.56 ± 0.15<br>(NS)<br>0.526 | 0.50 ± 0.21<br>(*)<br>0.042 | - | 0.91 ± 0.07<br>(**)<br>0.004 |
| Controls<br>(n = 53) | 0.88 ± 0.03<br>(****)<br>$1.62 \times 10^{-10}$ | 0.97 ± 0.07<br>(****)<br>$5.06 \times 10^{-6}$ | 0.91 ± 0.07<br>(**)<br>0.004 | - |

**Extended Data Tab. 2:**
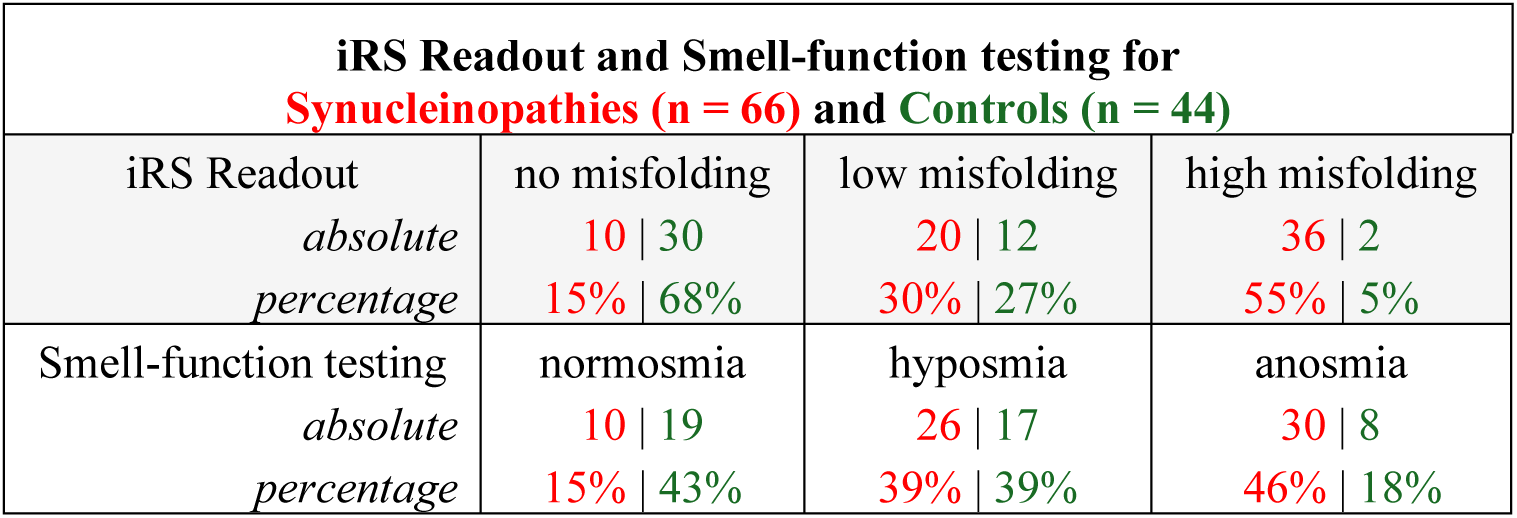
Comparison of iRS readout and smell-function testing in synucleinopathies and controls. Analysis of the iRS and smell-function test results in synucleinopathy and control groups. Results are shown for 110 individuals, including synucleinopathies (n = 66, red) and controls (n = 44, green), with values reported as absolute numbers and percentages. Among individuals with synucleinopathies, 55% were classified as high αSyn misfolding, 30% as low αSyn misfolding and 15% as no αSyn misfolding based on the iRS readout, whereas smell-function testing classified 46% as anosmic, 39% as hyposmic and 15% as normosmic. In the control group, 68% were classified as no misfolding, 27% as low misfolding and 5% as high misfolding by the iRS. Smell-function testing classified 43% of controls as normosmic, 39% as hyposmic and 18% as anosmic. Abbreviations: αSyn alpha-synuclein, iRS immuno-infrared-sensor

