## Supporting Information for "MISFOLDING OF ALPHA-SYNUCLEIN AS BLOOD-BASED BIOMARKER FOR PARKINSON’S DISEASE"

| Appendix Fig. | Description |
|---|---|
| S1 | Dataset distribution and conformity to normal distribution by normal Q-Q-Plot of spectral ratios. |
| S2 | Logistic regression models for the combined dataset. |
| S3 | iRS reproducibility measurement. |

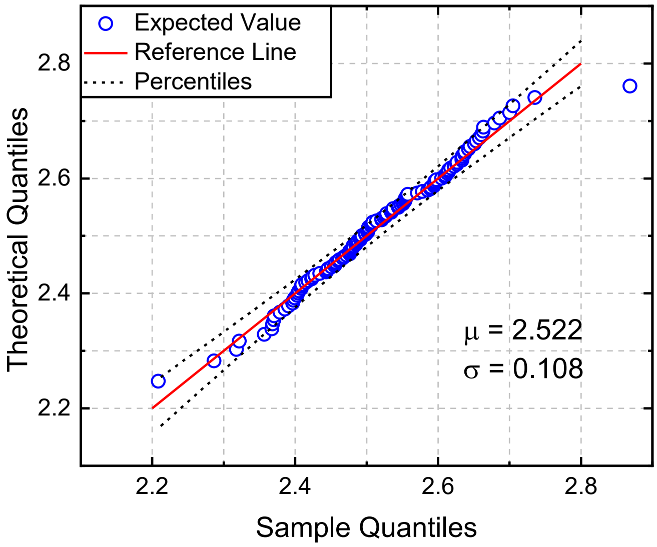


**Supplementary Fig. 1: Dataset distribution and conformity to normal distribution by normal Q-Q-Plot of combined ratio.**
Expected values are shown in blue dots, whereas the red line marks the reference line with 95% confidence bands (dashed black lines). The mean value of the normal distribution is $\mu = 2.522$ (SD, $\sigma = 0.108$). Typically, s-shaped expected values indicate a non-normal distribution, wherefore non-parametric significance tests were applied. Abbreviations: iRS immuno-infrared-sensor, SD standard deviation

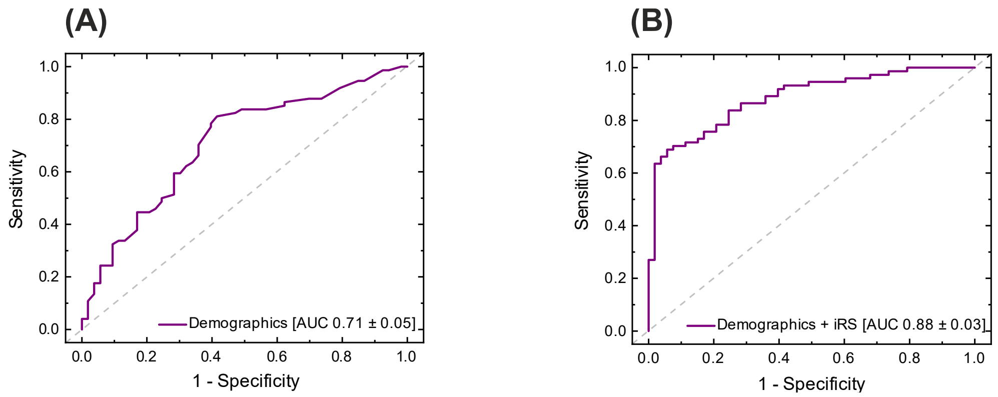


**Supplementary Fig. 2: Logistic regression models for the combined dataset.**

To assess diagnostic performance, two logistic regression models are applied with the clinical diagnosis as target variable. The first model ("Demographics") includes age and sex as independent variables, while the second model ("Demographics + iRS") additionally incorporates the iRS readout using the combined spectral ratio. ROC AUC values are 0.71 ± 0.05 for the demographics-only model and 0.88 ± 0.03 for the model including the iRS readout, demonstrating the added diagnostic value of iRS measurements.

Abbreviations: AUC area under the curve, iRS immuno-infrared-sensor, ROC AUC receiver operating characteristic

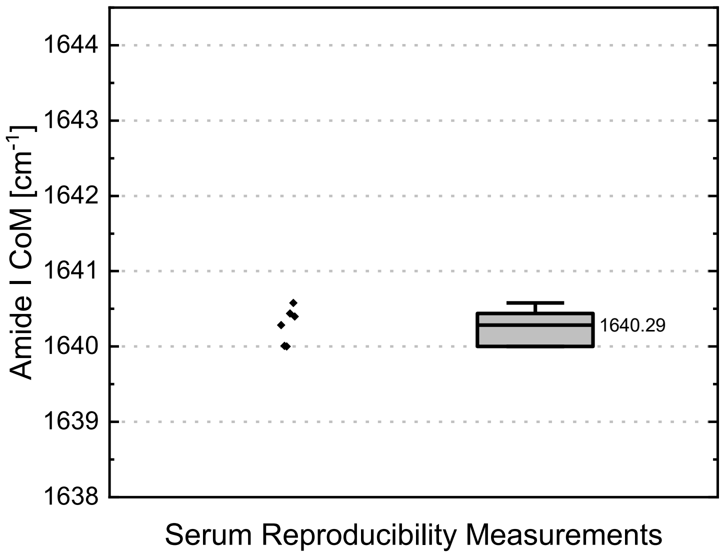


**Supplementary Fig. 3: Reproducibility measurements using a HC serum sample measured on the antibody-functionalized iRS surface.**

The same serum sample is measured in five independent runs to evaluate reproducibility. The amide I CoM, calculated from the upper 10–20% of the amide I band, is used as readout to enhance robustness. Spectral processing includes standard water vapor and baseline correction, FSD and smoothing. The mean CoM across replicates is 1640.29 $cm^{-1}$ with a standard deviation of 0.27 $cm^{-1}$. Box and whisker plots display the median (vertical line), interquartile range (boxes) and ± 1 SD (whiskers), with minimum and maximum values ranging from 1640.01 to 1640.44 $cm^{-1}$. Each point represents the iRS result from one measurement.

Abbreviations: CoM center of mass, FSD Fourier self-deconvolution, HC healthy control, SD standard deviation
